# Risk of Nephrolithiasis in adults heterozygous for *SLC34A3* Ser192Leu in an unselected health system cohort

**DOI:** 10.1101/2023.01.21.23284856

**Authors:** Chinedu Nwachukwu, Gurmukteshwar Singh, Bryn Moore, Natasha T Strande, Ion D Bucaloiu, Alexander R Chang

## Abstract

Homozygous or compound heterozygous mutations in solute carrier family 34, member 3 (*SLC34A3*) cause hereditary hypophosphatemic rickets with hypercalciuria (HHRH). Patients heterozygous for *SLC34A3* pathogenic variants may be at increased risk for renal calcification but reports have been mostly limited to family members of patients with autosomal recessive HHRH. To determine the phenotypic spectrum of *SLC34A3* Ser192Leu, we examined the most pathogenic *SLC34A3* variant Ser192Leu (238 out of 174,417 participants) in an unselected, health system-based research cohort in central and northeast Pennsylvania. *SLC34A3* Ser192Leu heterozygotes had higher risks of nephrolithiasis ICD diagnosis (13% vs. 6%), hypophosphatemia <2.5 mg/dL (31/96 [32%] vs. 6226/39636 [16%]), lower eGFR (−4.43, 95% CI: −7.03, −1.83; p=0.001) and tended to have higher prevalence of kidney/liver cyst ICD codes (5% vs. 3%; p=0.09), compared to controls. Further studies are needed to determine whether personalized approaches (i.e. phosphate supplementation) to patients heterozygous for *SLC34A3* pathogenic variants can reduce kidney stone burden and risk of kidney function decline.

## Introduction

Homozygous or compound heterozygous mutations in solute carrier family 34, member 3 (*SLC34A3*), encoding the sodium dependent inorganic phosphate cotransport proteins 2c (NPT2c), are known to cause autosomal recessive hypophosphatemic rickets with hypercalciuria (HHRH)(1) (2). This syndrome is characterized by hypophosphatemia, elevated 1,25-dihydroxyvitamin D (1,25(OH)_2_D), hypercalciuria and rickets/osteomalacia(2). Previous reports on patients with heterozygous pathogenic variants in *SLC34A3* have been mostly limited to family members of patients with autosomal recessive HHRH. Thus, the prevalence, penetrance, and phenotypic severity of *SLC34A3*-related disease is unknown(3). In this study we aimed to describe the prevalence and phenotypic spectrum of *SLC34A3* Ser192Leu, the most common pathogenic *SLC34A3* variant in non-Finnish European populations (allele frequency 0.001) (4) in an unselected, health system-based cohort in central and northeast Pennsylvania.

## Methods

The ethics committee/IRB of Geisinger Medical Center gave ethical approval for this work, and participants completed informed consent in the Geisinger MyCode/DiscovEHR study (5). We identified patients with *SLC34A3* Ser192Leu variants and extracted linked EHR data (demographics, pertinent labs and International Classification of Diseases [ICD] diagnosis codes). The primary outcome was nephrolithiasis; secondary outcomes included outpatient hypophosphatemia (lowest phosphate <2.5 mg/dL; median phosphate <3.0 mg/dL; hypophosphatemia meeting both criteria), and median phosphate and last eGFR as continuous variables. We also examined a secondary outcome of any kidney/liver cyst, which included ICD codes for ADPKD as well as cystic kidney diseases, congenital kidney cyst, or liver cystic disease. Chart review was performed by a nephrology fellow (CN), focused on nephrolithiasis and nephrocalcinosis diagnosis and management, nephrology and urology visits, imaging reports, 24-hour urine stone risk profiles, relevant medications (calcium, phosphorus, vitamin D supplements, thiazide diuretics), and urologic procedures. EHR-based renal phenotypes were compared between *SLC34A3* Ser192Leu heterozygotes and controls (without any *SLC34A3* variants) using logistic and linear regression for categorical and continuous outcomes, respectively, adjusted for age, sex, genetic ancestry, and clustered by family network. An exploratory analysis examining the relationship between serum phosphate and 24-hour urine calcium in stone-formers with available data was conducted. STATA/MP 15.1 (College Station, TX) was used for analyses.

## Results and discussion

Out of 174,417 participants, we identified 238 individuals (136 per 100,000) with the *SLC34A3* Ser192Leu variant. We excluded 3 individuals who had an additional *SLC34A3* variant of uncertain significance and 17 missing EHR data. Demographic and clinical characteristics of 218 individuals heterozygous for the Ser192Leu variant are presented in **Table**. In brief, there were 136 (62%) females, of mean (SD) age 58.6 (18.6) years, and the vast majority (99%) were of European ancestry. *SLC34A3* Ser192Leu heterozygotes were at increased risk of nephrolithiasis ICD diagnosis (13% vs. 6%), minimum phosphate <2.5 mg/dL (31/96 [32%] vs. 6226/39636 [16%]), median phosphate <3.0 mg/dL (27/96 [28%] vs. 5517/37983 [15%]), and hypophosphatemia meeting both criteria (21/96 [22%]) vs. 2994/37983 [8%]) (p<0.001 for all comparisons). *SLC34A3* Ser192Leu had lower eGFR (−4.43, 95% CI: −7.03, −1.83; p=0.001) and prevalent eGFR <60 ml/min/1.73m^2^ (52/190 [27%] vs. 30255/148784 [20%] (p=0.04) and tended to have higher prevalence of kidney/liver cyst ICD codes (5% vs. 3%; p=0.09), compared to controls.

**Table.** Characteristics of *SLC34A3* Ser192Leu heterozygotes and controls without *SLC34A3* variants.

| Table. Characteristics of *SLC34A3* Ser192Leu heterozygotes and controls without *SLC34A3* variants |  |  | aOR* (95% CI) |
| --- | --- | --- | --- |
|  | *SLC34A3* Ser192Leu heterozygotes (n=218) | Controls (n=168,882) |  |
| Follow-up time, y (SD) | 14.72 (5.49) | 13.85 (6.00) |  |
| Age, y (SD) | 58.64 (18.56) | 57.36 (18.91) |  |
| Female, % | 136 (62) | 101952 (61) |  |
| European Ancestry, % | 213 (99) | 155440 (94) |  |
| eGFR available, % | 190 (87) | 148628 (88) |  |
| eGFR, ml/min/1.73m<sup>2</sup> | 74.23 (23.55) | 80.91 (26.70) |  |
| Serum phosphate available, % | 96 (44) | 39636 (23) |  |
| Minimum serum phosphate, mg/dL | 2.76 (0.73) | 3.08 (0.69) |  |
| Median serum phosphate, mg/dL | 3.17 (0.66) | 3.43 (0.61) |  |
| Nephrolithiasis ICD code, % | 28 (13) | 9767 (6) | 2.38 (1.59, 3.56);<br>p<0.001 |
| Minimum phosphate <2.5 mg/dL, % | 31/96 (32) | 6226/39636 (16) | 2.67 (1.74, 4.11);<br>p<0.001 |
| Median phosphate <3.0 mg/dL, % | 27/96 (28) | 5768/39636 (15) | 2.42 (1.52, 3.86);<br>p<0.001 |
| Both minimum phosphate <2.5 and median phosphate <3.0 mg/dL, % | 21/96 (22) | 3125/39636 (8) | 3.41 (2.05, 5.68);<br>p<0.001 |
| **eGFR <60, ml/min/1.73m<sup>2</sup>** | 52/190 (27) | 30255/148784 (20) | 1.45 (1.01, 2.07); p=0.04 |
| **Kidney/Liver cysts, %** | 11/218 (5) | 5084/168882 (3) | 1.68, 95% CI: 0.92, 3.08<br>p=0.09 |

ICD 9/10 codes used were as follows: nephrolithiasis (N20, 592.0, 592.1, 592.9, 594.0, 594.1, 594.2, 594.8, 594.9, 274.11), kidney/liver cysts (Q61.2, Q61.3, 753.13, 753.12, 753.10, Q61.5, Q61.8, Q61.9, Q61.00, Q61.01, Q61.02, 753.11, 753.19, Q44.6, 573.8, 751.62)
\*Models adjusted for age, sex, genetic ancestry, clustered by family network.
Abbreviations: aOR (adjusted odds ratio), ICD (International Classification of Diseases), eGFR (estimated glomerular filtration rate)

In the subset of 9 stone formers with 24-hour urine data, 6 (67%) had hypercalciuria (urine calcium excretion > 250 mg/24h in men and > 200 mg/24h in women), and 3 (33%) had hyperphosphaturia (urine phosphorus excretion > 1100 mg/24h). Three patients underwent analysis of kidney stones; all were calcium oxalate. Minimum phosphate level was strongly correlated with 24-hour urine calcium (r=−0.76; p=0.03) (**Figure**).

**Figure.**
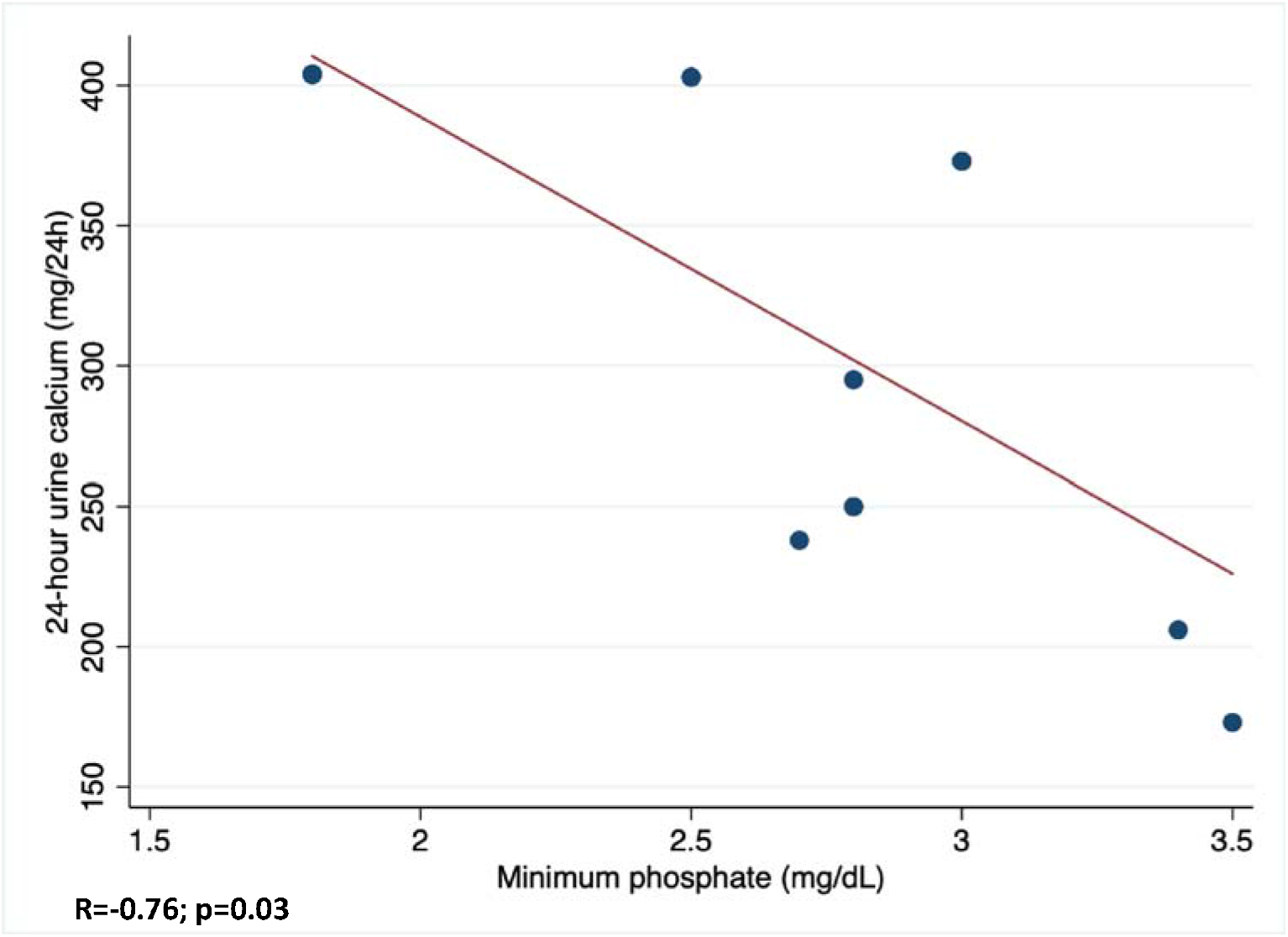
Relationship between Serum Phosphate with 24-hour Urine Calcium among 8 Kidney stone formers. Among 8 *SLC34A3* Ser192Leu heterozygotes with nephrolithiasis and available data, minimum serum phosphate levels correlate strongly with 24-hour urine calcium levels (R=−0.76; p=0.03).

Focused chart review revealed additional history of nephrolithiasis not captured by ICD code with a total of 66 (30%) having evidence of nephrolithiasis, including 13 on imaging only. Only 2 (0.9%) had evidence of nephrocalcinosis on imaging available in the EHR. Eleven (5%) had a family history of nephrolithiasis. None of the 66 affected individuals with nephrolithiasis had undergone clinical genetic testing for nephrolithiasis. Twenty-one (32%) patients were seen by urology, and 20 (30%) patients underwent urologic procedures. Only 13 (20%) stone formers received stone prevention evaluation in nephrology. Review of medications revealed that 12 (6%) patients were receiving calcium supplements, and 29 (13%) patients were receiving vitamin D supplements.

In this unselected health system-based cohort, we found that *SLC34A3* Ser192Leu heterozygotes had ∼2-3-fold higher risks of nephrolithiasis and hypophosphatemia. Our findings are consistent with another cohort of 120 (61 monoallelic, 43 biallelic, 16 wild-type) individuals from 27 families with *SLC34A3* pathogenic variants and available data (3). Dasgupta et al. reported that individuals with biallelic mutations in *SLC34A3* alleles had an approximately tenfold higher risk of nephrolithiasis and nephrocalcinosis whereas heterozygotes had approximately 3-fold higher risk of nephrolithiasis (16.4%) compared with the general population (3). Another recent case series of 12 patients with *SLC34A3* pathogenic variants (7 monoallelic, 5 biallelic) described a higher preponderance for kidney cysts than the general population(7). In our study *SLC34A3* Ser192Leu heterozygotes had numerically more kidney/liver cyst ICD codes though this result was not statistically significant. In addition, our study found a borderline association with decreased eGFR <60 ml/min/1.73m^2^. A limitation of this study was the lack of standardized imaging and biochemical measurements on all participants. The strengths of our study included an unselected health system-based cohort with long duration of follow-up allowing us to more fully capture the burden of nephrolithiasis among this population.

This study has potentially important implications for patients heterozygous for *SLC34A3* pathogenic variants. Very few received 24-hour urine stone risk profile testing, making it very difficult to identify hypercalciuria and hyperphosphaturia. Many patients were receiving calcium and vitamin D supplements, which would be expected to worsen nephrolithiasis risk. Individuals with biallelic *SLC34A3* variants are known to have renal phosphate wasting, elevated calcitriol levels, and hypercalciuria, which reverse with oral phosphorus supplementation(1) (6).

Heterozygotes have an intermediate phenotype with borderline-low serum phosphate levels, which stimulates calcitriol synthesis, and resultant hypercalciuria (3). Knowledge of a *SLC34A3* pathogenic variant may enable clinicians to treat hypophosphatemia with oral phosphate supplementation to offset the stimulus for increased calcitriol synthesis. Further studies are needed to determine whether a precision medicine approach to nephrolithiasis identifying pathogenic heterozygous variants in *SLC34A3* can reduce kidney stone burden and risk of kidney function decline in this population at high risk for nephrolithiasis.

## Data Availability

All data produced in the present work are available upon reasonable request to the authors and subject to a data use agreement.

## Acknowledgements

We thank the participants of the MyCode Community Health Initiative and the Geisinger-Regeneron DiscovEHR Collaboration for use of the genomic and electronic health information. We are also grateful for Thomas Jones for assistance with extracting EHR data and Tooraj Mirshahi and Jonathan Luo for the use of the genetic database.

